# Experiments and Simulations to Assess Exercise-Induced Pressure Drop Across Aortic Coarctations

**DOI:** 10.1101/2024.11.14.24317284

**Authors:** Priya J. Nair, Emanuele Perra, Doff B. McElhinney, Alison L. Marsden, Daniel B. Ennis, Seraina A. Dual

## Abstract

Blood pressure gradient (ΔP) across an aortic coarctation (CoA) is an important measurement to diagnose CoA severity and guide treatment. While invasive cardiac catheterization is the clinical gold-standard for measuring ΔP, it requires anesthesia and does not capture the effects of daily activity or exercise, potentially underestimating the disease’s functional burden. This study aimed to identify patients with functionally significant CoA by evaluating exercise-induced ΔP using a hybrid mock circulatory loop (HMCL). Patient-specific aorta geometries (N=5) of patients with CoA were generated from 4D-Flow magnetic resonance imaging (MRI) scans, then 3D-printed to create compliant aortic phantoms. The phantoms were incorporated into an HMCL with flow and pressure waveforms tuned to patient-specific rest and exercise states. Matched fluid-structure interaction (FSI) simulations were performed using SimVascular for comparison. Results showed that mean ΔP increased non-linearly with cardiac output (CO), with trends differing between patients. HMCL and FSI simulations exhibited excellent agreement in trends of ΔP change with CO, with minimal error of 1.6 ± 1.1 mmHg. This study emphasizes the need for assessing exercise CoA hemodynamics beyond resting ΔP measurements. Overall, HMCLs and FSI simulations enable assessment of patient-specific hemodynamic response to exercise unattainable in clinical practice, thereby facilitating a comprehensive non-invasive assessment of CoA severity. Further, the excellent agreement between HMCL and FSI results indicates that our validated FSI approach can be used independently to assess exercise CoA hemodynamics hereafter, eliminating the need for repeated complex HMCL experiments.

## 1 Introduction

Coarctation of the aorta (CoA) is a congenital heart disease characterized by a constriction of the aorta, typically just distal to the origin of the left subclavian artery. CoA accounts for 5-7% of all congenital heart diseases [1], with an incidence of approximately 3 cases per 10,000 births [2]. The narrowing results in increased resistance to blood flow, hypertension in the upper extremities, and an elevated pressure gradient (Δ P) across the CoA. If left untreated, CoA can lead to premature coronary artery disease, ventricular dysfunction, aortic aneurysms, aortic dissection, and cerebral vascular disease [3].

Current treatment methods include surgical correction and catheter-based stent implantation. In the current standard of care, ΔP at rest (measured during catheterization) is used to assess the severity of the CoA and to determine whether a patient should undergo corrective intervention. Since ΔP increases during exercise due to increased cardiac output (CO), patients with ΔP below the threshold for intervention at rest (20 mmHg at peak systole) may experience pathologically high ΔP during moderate exercise, although this response varies on a patient-specific basis [4]. In such cases, the catheterization at rest underestimates the functional disease burden in everyday life. In the clinical setting, adrenergic drug (dobutamine) infusion can be used during catheterization to simulate exercise and provoke an exercise-induced pressure gradient, but studies have shown that the hemodynamic response during pharmacological stress does not accurately represent the responses to physical exercise [5,6].

The complexity of the cardiovascular system as well as the variability in physiology and presentation of disease make it difficult to select appropriate treatment methods and identify patients at risk. Mock circulatory loops (MCLs) can provide a mechanical platform for replicating specific physiological or pathological hemodynamics on the benchtop [7–9]. In a typical MCL, hydraulic components such as pumps, reservoirs, and pipes are physically tuned to mimic resistance to blood flow, compliance of the vessel walls, and blood’s inertia, therefore creating an *in vitro* representation of the cardiovascular system [8,10,11]. However, one major limitation of such MCLs is the difficulty in tuning physical parameters (e.g. clamp forces, air reservoir pressure, fluid volumes). Hybrid MCLs (HMCLs) take this a step further and allow extra flexibility and rigor by incorporating a combination of *in vitro* and *in silico* elements [12–14]. In HMCLs, the phantoms interface with a computational closed-loop lumped-parameter network (LPN) representation of the cardiovascular system tuned to match patient-specific hemodynamics. The patient-specific LPN of the HMCL eliminates the complexity of physical tuning of resistance and compliance elements compared to standard MCLs while achieving patient-specific flows and pressures. Prior studies have used HMCLs for a variety of applications ranging from studying hemodynamics in Fontan graft obstruction to evaluating the performance of total artificial hearts [12,15–17]. The use of HMCLs to guide clinical decision-making in patients with CoA is only explored in standard geometries [14]. The advantage of this approach lies in its ability to replicate patient-specific hemodynamic states otherwise unattainable in clinical practice for the evaluation of ΔP.

Fluid-structure interaction (FSI) simulations have been used extensively to non-invasively assess local hemodynamics in patients with cardiovascular disease [18–21]. Anatomical and physiological data from routine clinical imaging can be used to create patientspecific geometries and simulate hemodynamics. Our previous work has shown the advantages of using reduced-order (0D) models to not only tune boundary conditions, but also to initialize 3D simulations [22]. These computationally efficient models can be used to speed up some of the time-consuming steps in setting up and running a 3D simulation.

The objective of this study was to identify patients with functionally significant CoA by measuring exercise-induced pressure drop in compliant phantoms of CoA using a patient-specific tuned hybrid mock circulatory loop (HMCL) *in vitro*. Additionally, we compare our *in vitro* findings with results from patient-specific FSI simulations. Our study provides experimental and simulation evidence of non-linear effects on ΔP evident during moderate exercise in patients with CoA.

## 2 Methods

In this section, we summarize the methods used to create patientspecific compliant aortic phantoms and to evaluate hemodynamics at rest and exercise using an HMCL. We also outline the FSI simulations approach, which we used to compare to the measurements obtained from the HMCL.

### 2.1 Patient Data Acquisition

Under a protocol approved by the Stanford Institutional Review Board (IRB), five patients with CoA from the Lucile Packard Children’s Hospital at Stanford were retrospectively identified. Patients with native and recurrent CoA were included. Inclusion criteria for the cohort required a 4D-Flow MRI exam and invasive blood pressure (BP) measurements acquired via cardiac catheterization. We obtained retrospective 4D-Flow MRI datasets for five patients with CoA (Table 1, age = 31 ± 16.8 years, 4M / 1F). Invasive measurements of pressure via cardiac catheterization in the ascending aorta (AAo) and descending aorta (DAo) were obtained, in addition to heart rate (HR) and cardiac index (CI). Informed consent was not required for this retrospective clinical data collection. Using the available information, systemic vascular resistance (*SV R*) and pulmonary vascular resistance (*PV R*) were calculated as follows:

**Table 1.** Patient characteristics. BSA: Body surface area, CO: Cardiac output, HR: Heart rate.

| Patient | Sex | BSA (m<sup>2</sup>) | CO (L/min) | HR (bpm) |
| --- | --- | --- | --- | --- |
| P-1 | M | 1.94 | 8.0 | 71 |
| P-2 | F | 1.56 | 4.4 | 71 |
| P-3 | M | 1.58 | 3.6 | 89 |
| P-4 | M | 1.89 | 5.5 | 62 |
| P-5 | M | 2.09 | 6.5 | 71 |

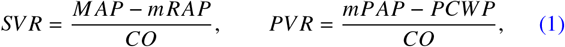

where *M AP* is the mean arterial pressure, *mR AP* is the mean right atrial pressure, *mPAP* is the mean pulmonary arterial pressure, *PCW P* is the pulmonary capillary wedge pressure, and *CO* is the cardiac output.

### 2.2 Patient-specific Phantom Generation

MRI magnitude images were imported into SimVascular (simvascular.org) [23]. Centerlines were manually drawn through the aorta and the branches arising from the aortic arch: the brachiocephalic trunk, the left common carotid artery, and the left subclavian artery. 2D contours were manually drawn along the centerlines to trace the blood vessel lumen and then lofted to produce a patient-specific model of the blood volume. We used Meshmixer (Autodesk, San Francisco, CA, USA) to generate the outer wall (wall thickness = 2 mm) and to add cylindrical caps (length = 20 mm) at the inlets and outlets to enable connection to customized barbed transition elements that connect the phantom to tubing. We used a photopoly-merization 3D printer (J735 PolyJet, Stratasys, Eden Prairie, MN, USA) with a compliant printing material (Agilus30, Stratasys) to manufacture compliant wall phantoms of the patient-specific aorta geometries (Figure 1) with an elastic modulus matching that of a typical human aorta (Y = 1.2 MPa) [24,25]. The printed phantoms were coated with a thin conformal coating (DOWSIL 3-1953) to prevent fluid absorption.

**Fig. 1.**
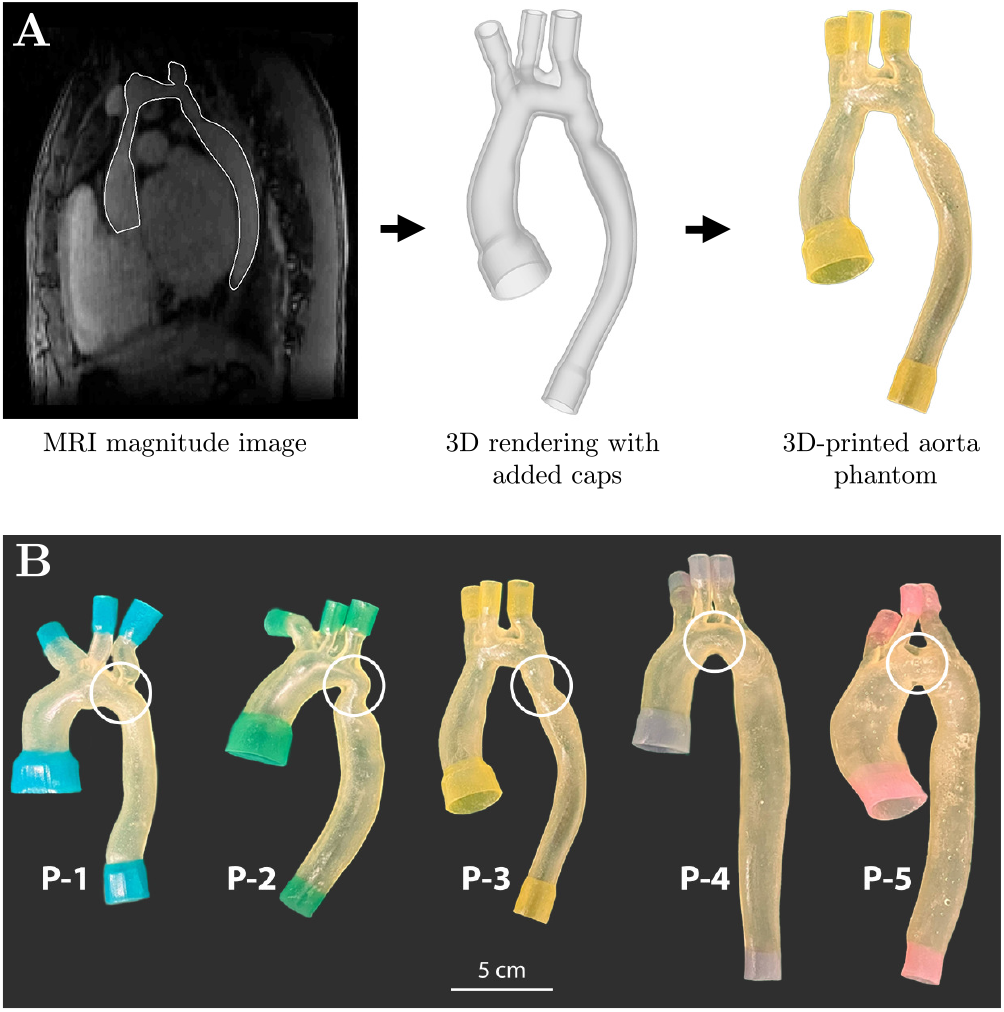
(A) Patient-specific phantom generation process, (B) 3D-printed patient-specific phantoms of CoA with compliant walls. White circles indicate location of CoA.

### 2.3 Hybrid Mock Circulatory Loop (HMCL)

We assessed the hemodynamics in the aortic phantoms at rest and exercise condition using an HMCL (Figure 2 [12,14]. HMCLs incorporate elements of both *in silico* and *in vitro* modeling allowing for testing of physical elements in interaction with a dynamic closed-loop cardiovascular LPN. The compliant aortic phantoms were physically integrated into the HMCL *in vitro* while the remainder of the cardiovascular system was represented through a closed-loop cardiovascular LPN *in silico*. A graphical representation of the HMCL setup is shown in Figure 2A. p_1_, p_2_, and p_3_ are the pressures in Tanks-1, 2, and 3, respectively, as simulated in the cardiovascular LPN and imposed on the tanks. q_1_ and q_2_ are the flows measured at the AAo inlet and DAo outlet, respectively. The components of the HMCL are described in further detail below.

**Fig. 2.**
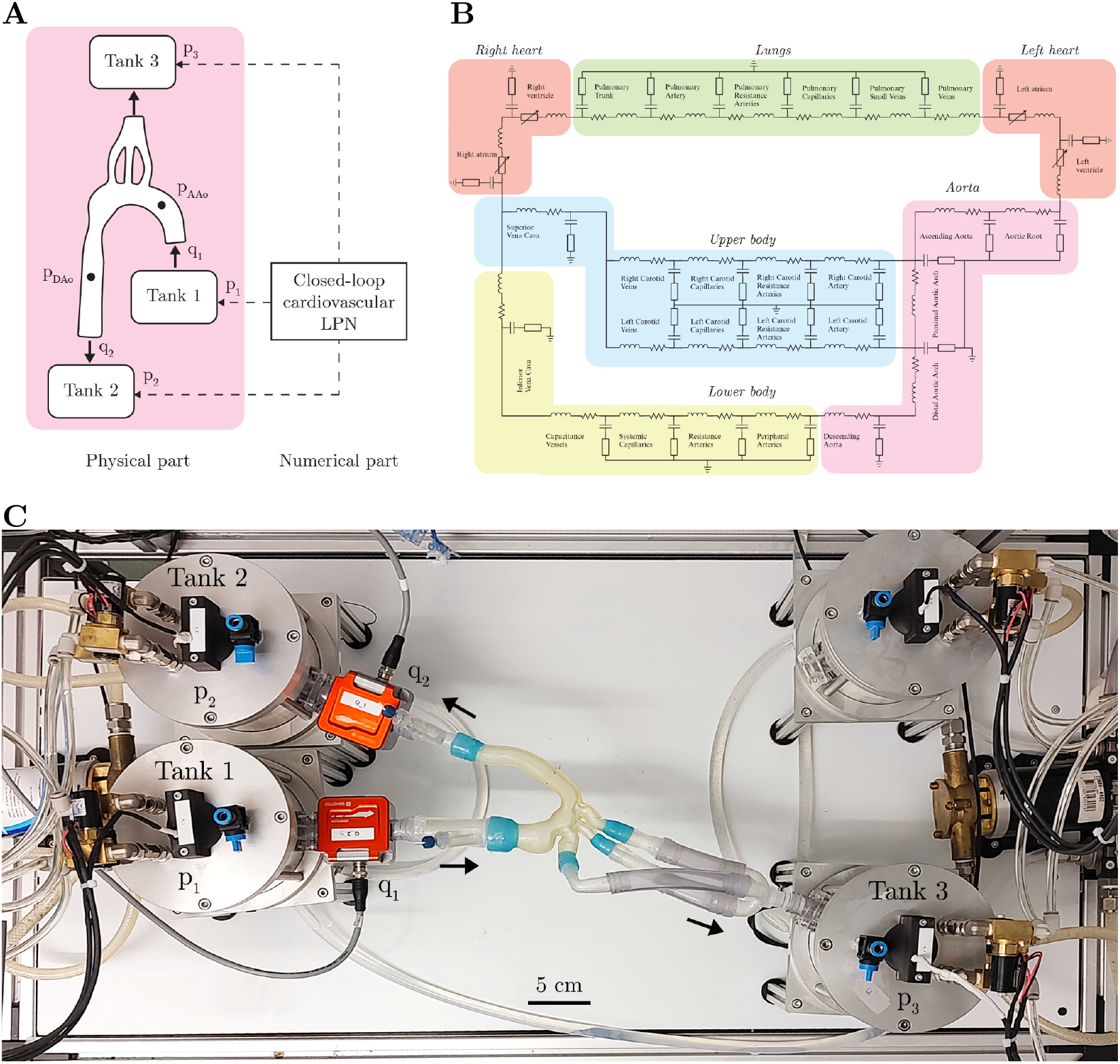
(A) HMCL setup schematic. (B) Closed-loop LPN model of the cardiovascular system. (C) *In vitro* components of the HMCL. Black arrows indicate the direction of flow. Flow sensors are shown in red.

#### 2.3.1 In vitro: Closed-loop cardiovascular LPN

The closedloop LPN of the cardiovascular system used in this study (Figure 2B) is based on a model developed by Broomé et al. [26]. The LPN model includes 25 vascular compartments, each representing a specific section of the systemic and pulmonary circulation (Figure 2B). Patient-specific physical dimensions (length, radius, wall thickness) of the aortic arch compartments (aortic root, ascending aorta, proximal aortic arch, right and left carotid artery, distal aortic arch) are used as input parameters to the vascular phantom. Each vasculature segment is modeled as a 4-element Windkessel model. Depending on the severity of the CoA, flow non-linearities may emerge at the stenosed vessel location, making the Poiseuille-flow assumption invalid. For this reason, the coarctation element is modelled as a non-linear expansion of the in-series resistance of the 4-element Windkessel model [27]. The heart is modeled with four distinct chambers, with their active and passive properties described using a periodic “double-Hill” function [28]. Cardiac valves are modeled using a combination of a Bernoulli resistance and an inertial term, allowing the valve areas to change gradually as a function of pressure gradient [29]. The closed-loop cardiovascular LPN also accounts for intra-pericardial pressure [30] and atrioventricular septal displacement [31]. The closed-loop cardiovascular LPN is implemented in MATLAB Simulink (MathWorks Inc., Natick, MA, USA) and run as a real-time system through the MicroLabBox (MicrolabBox, dSPACE GmbH, Paderborn, Germany) at a sampling frequency of 2000 Hz.

#### 2.3.2 Simulation of rest and exercise

We numerically simulated *in vitro* three to five patient-specific hemodynamic states between rest and exercise in the closed-loop cardiovascular LPN, defined by different levels of CO: 0.75×CO, rest (1.0×CO), 1.25 CO, 1.5 CO, and 2×CO. For the resting state, the target HR, SVR, and PVR obtained from the catheterization report were used as input parameters. In order to make the cardiovascular LPN patientspecific, we adjusted the Young’s modulus of the vessel walls in the systemic and pulmonary arteries, along with the maximum elastance of both ventricles, to align with the patient’s resting CO and match the systolic and diastolic pressures (SBP, DBP) within a 5 mmHg tolerance. Clinical measurements of HR, SVR, and PVR during exercise were not available. Target values of HR, SVR and PVR during exercise states were derived from existing literature. For a 50% increase in CO with exercise, HR was increased by 50%, SVR decreased by 20%, and PVR decreased by 15% from rest in accordance with a previous study performed in patients with CoA undergoing MRI-ergometry [32]. We obtained the parameters for the other exercise states by linear interpolation/extrapolation. For initialization, the closed-loop LPN was first tuned as a numerical standalone model before connecting it to the physical components of the HMCL where it was fine-tuned.

### 2.3.3 In vitro Hardware Components

The pressure-controlled fluid reservoirs (tanks) are the main hardware components of the HMCL. They form the physical interface between the closed-loop cardiovascular LPN and the aortic phantom by converting the simulated pressure waveforms into controlled pressure actuation with real-time precision. The fluid pressure in the tanks is controlled by adjusting the pressure of the air cavity in the tanks through exchange of pressurized air or vacuum. The fluid pressure is constantly monitored by pressure sensors (PN2099, IFM Electronic

Geräte GmbH & Co. KG, Essen, Germany) placed at the bottom of each reservoir. The compliant aortic phantoms are connected to the tanks via customized 3D-printed connectors and PVC tubing in the configuration shown in Figure 2C. Tank-1 is connected to the aorta inlet, Tank-2 to the DAo outlet, and Tank-3 to the head and neck vessel outlets. Real-time measurements from ultrasonic flow probes (Sonoflow CO.55/190, Sonotec GmbH, Halle, Germany) that measure flow in the phantom drive the backflow pump to generate unidirectional flow from Tanks-2 and 3 back to Tank-1 to minimize the difference in fluid level between the tanks. The working fluid was a mixture of water and glycerol (volume ratio 5:3) with viscosity matching that of blood. We used pressure transducers (SPR-350S, Millar) to measure invasive BP. Catheters were inserted in the phantom through Tuohy-Borst adapters to measure local pressure at the AAo and DAo over the cardiac cycle. The flow probes provided flow measurements at the AAo and DAo. Once steady state was reached, we recorded measurements for one minute. To reduce noise, we filtered the catheter signals using a low-pass filter with a patient-specific cut-off frequency between 5 and 10 Hz. Mean pressure drop (ΔP) was estimated as the difference in mean pressure between AAo and DAo. We report the average of the mean pressure drop (ΔP_HMCL_) across 5 cardiac cycles.

### 2.4 FSI Simulations

We compared ΔP measurements acquired from the HMCL against those from 3D FSI simulations. The inflow waveform measured in the HMCL was prescribed as the inlet boundary condition of the FSI simulation. A three-element Windkessel model (proximal resistance *R*_p_, capacitance *C*, distal resistance *R*_d_) was imposed at each of the outlets in an open-loop fashion. At rest, the total peripheral resistance (*R*_total_) was determined using mean inflow and mean aortic BP (*P*_mean_) measured in the HMCL as follows:

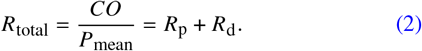

Capacitance *C* and resistance ratio *R*_p_:*R*_d_ were iteratively tuned to match flow splits and pressures achieved in the HMCL within 5 mmHg using the approach outlined in Nair *et al*. [22]. For each of the exercise states, *R*_total_ was reduced by the same percentage that *SV R* was reduced in the HMCL, then divided into *R*_p_ and *R*_d_. The tuned boundary conditions are then applied to the 3D FSI model. We initialized all the 3D simulations with tuned 0D simulations, thereby allowing the 3D simulations to converge faster than they would have without any initialization [22,33]. We performed 3D simulations using the coupled momentum method (CMM) with svSolver, SimVascular’s finite element solver for fluid-structure interaction between an incompressible, Newtonian fluid and a linear elastic membrane for the vascular wall [23,34]. The Young’s modulus of the vessel wall was prescribed to be 1.2 MPa based on tensile testing performed previously on the 3D-printed material of the aortic phantoms [25]. The thickness of the wall was assumed to be 10% of the diameter of the wall [19,35]. A Poisson’s ratio 0.5 and density 1 g/cm^3^ were used to further define the vessel wall material properties. The fluid was prescribed to have density 1.06 g/cm^3^ and viscosity 0.04 Poise to match the properties of blood. 3D simulations were run for 10 cardiac cycles to ensure that the pressures reach periodic convergence; only the final cardiac cycle was analyzed. We report the mean pressure drop between AAo and DAo (ΔP_FSI_) from the last cardiac cycle.

### 2.5 Statistical Analysis

The agreement between ΔP_HMCL_ and ΔP_FSI_ was characterized with Bland-Altman analysis. We performed bootstrapping to estimate the population mean absolute error and standard deviation of the population based on the sample of 5 patients with CoA.

## 3 Results

### 3.1 Achieving exercise in cardiovascular LPN

A total of five patients with CoA were included in this study. The parameters in the cardiovascular LPN of the HMCL were tuned to match clinical targets and to achieve rest and exercise in the patient-specific HMCL experiments. The patient-specific values at which the experiments were performed are listed in Table 2.

**Table 2.** HMCL parameters for rest and exercise. CO: Cardiac Output, HR: Heart Rate, SVR: Systemic Vascular Resistance, PVR: Pulmonary Vascular Resistance, BP: Blood Pressure in the AAo.

| Patient | State | CO<br>(L/min) | HR<br>(bpm) | SVR<br>(mmHg·s/mL) | PVR<br>(mmHg·s/mL) | BP (mmHg)<br>P<sub>sys</sub>/P<sub>dias</sub>/P<sub>mean</sub> |
| --- | --- | --- | --- | --- | --- | --- |
| P-1 | 0.75× | 5.96 | 53 | 0.67 | 0.065 | 112 / 59 / 73 |
|  | Rest | 7.61 | 71 | 0.59 | 0.058 | 118 / 69 / 82 |
|  | 1.25× | 9.64 | 88 | 0.37 | 0.041 | 114 / 40 / 65 |
| P-2 | Rest | 4.38 | 71 | 0.87 | 0.16 | 112 / 40 / 67 |
|  | 1.25× | 5.58 | 88 | 0.78 | 0.15 | 120 / 44 / 74 |
|  | 1.5× | 6.55 | 100 | 0.72 | 0.139 | 133 / 46 / 80 |
|  | 2× | 8.67 | 130 | 0.57 | 0.117 | 133 / 47 / 84 |
| P-3 | Rest | 3.69 | 89 | 0.85 | 0.329 | 79 / 43 / 56 |
|  | 1.25× | 4.74 | 111 | 0.76 | 0.309 | 84 / 48 / 61 |
|  | 1.5× | 5.44 | 133 | 0.68 | 0.269 | 88 / 50 / 64 |
|  | 2× | 7.29 | 170 | 0.63 | 0.22 | 94 / 48 / 66 |
| P-4 | Rest | 5.36 | 62 | 0.58 | 0.089 | 93 / 36 / 56 |
|  | 1.25× | 6.93 | 77 | 0.51 | 0.083 | 98 / 39 / 60 |
|  | 1.5× | 8.42 | 93 | 0.46 | 0.080 | 105 / 41 / 64 |
|  | 2× | 10.03 | 120 | 0.33 | 0.059 | 108 / 48 / 69 |
| P-5 | Rest | 6.40 | 71 | 0.70 | 0.095 | 108 / 63 / 82 |
|  | 1.25× | 8.14 | 88 | 0.63 | 0.075 | 117 / 70 / 91 |
|  | 1.5× | 9.68 | 105 | 0.57 | 0.068 | 125 / 76 / 97 |

The errors between our tuned cardiovascular LPNs and the targets derived from patient-specific clinical measurements is shown in Figure 3A. We achieved the target CO, HR, SVR and PVR with a mean absolute error of 2.3%, 1.1%, 5.1%, and 4.1% respectively across all patients and hemodynamic states. Particularly, matching the target SVR at 2× CO while still achieving the target CO was challenging. Figure 3B shows the errors observed between output from the cardiovascular LPN connected to the HMCL and measurements from the HMCL. Although the total CO measured in the HMCL was matched to that computed in the cardiovascular LPN, the peak flow was not matched due to hardware restrictions (Figure 3). Therefore, instead of reporting the peak systolic ΔP measurement in this study, we report the mean ΔP between the AAo and DAo, where mean ΔP refers to the average pressure drop between the AAo and DAo over the cardiac cycle.

**Fig. 3.**
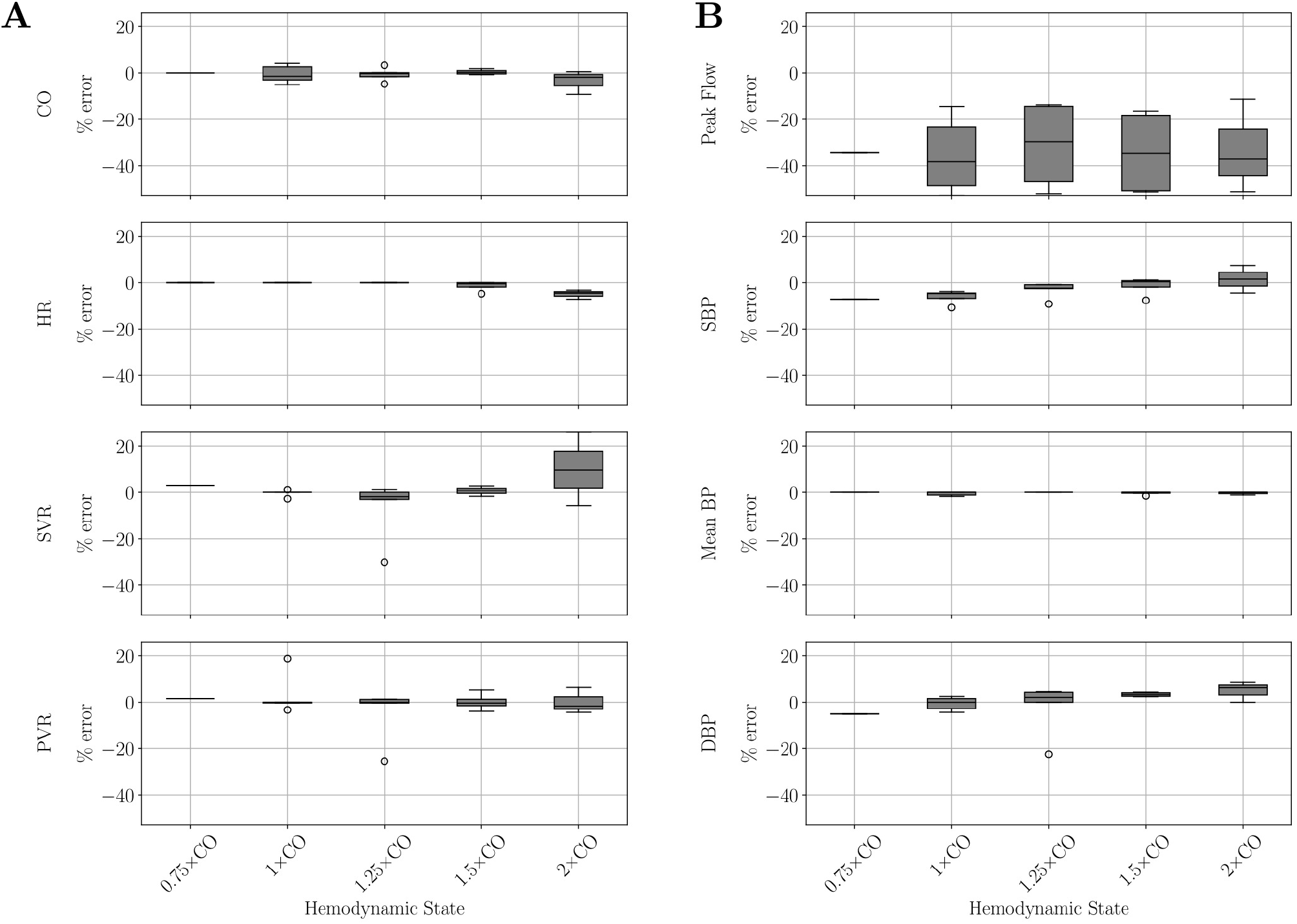
Percentage error between values measured in the HMCL and target values of eight HMCL parameters: CO, HR, SVR, PVR, peak flow, SBP, mean BP, and DBP at different hemodynamic states. (A) Target values are derived from clinical measurements and literature references for percentage changes with exercise. (B) Target values are output from the cardiovascular LPN connected to the HMCL. Median % error for each parameter across all patients at each hemodynamic state is represented by the black line. CO: Cardiac Output, HR: Heart Rate, SVR: Systemic Vascular Resistance, PVR: Pulmonary Vascular Resistance, SBP: Systolic Blood Pressure, DBP: Diastolic Blood Pressure.

Representative flow and pressure waveforms over a one second time interval at the AAo inlet in P-3 at different levels of exercise are shown in Figure 4. We observe that the increase in CO with increasing intensity of exercise is primarily due to an increase in HR (demonstrated by the increase in frequency in the top row of Figure 4A). Additionally, an increase in mean BP is observed with an increase in CO (Figure 4B), with the exception of P-1 at 1.25× CO (dotted line), a limitation that we address later.

**Fig. 4.**
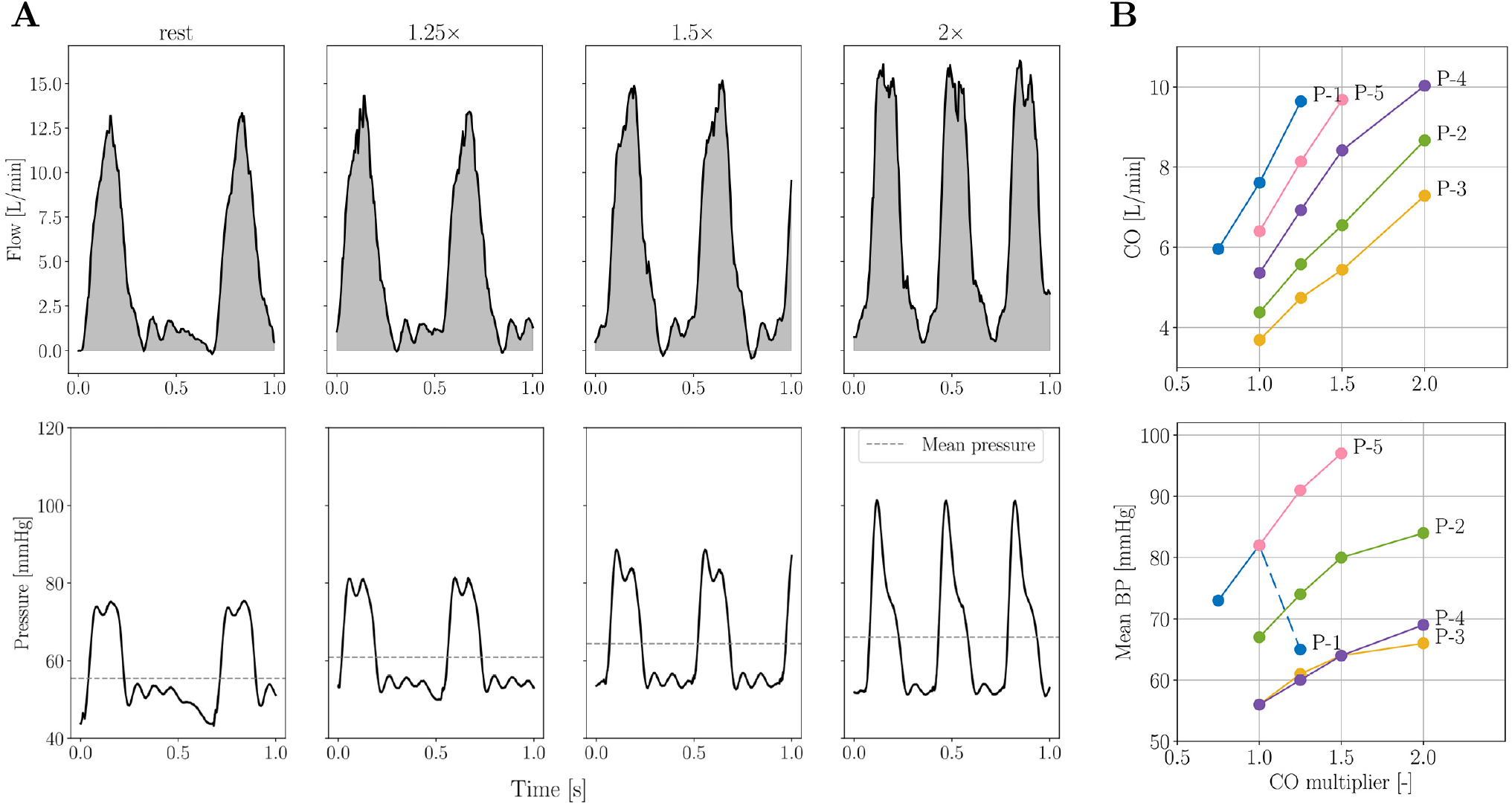
(A) Flow (top row) and pressure (bottom row) in the AAo and DAo at rest and different exercise states in P-3. (B) Increase in CO (top) and mean BP (bottom) with exercise.

### 3.2 Exercise-induced pressure drop

We observe that the increase in ΔP_HMCL_ is non-linear with the increase in CO and the response varies on a patient-specific basis. In some patients, such as P-4, ΔP_HMCL_ is relatively stable with only a 1.5 mmHg increase going from rest to 2× CO. On the other hand, in P-2, ΔP_HMCL_ increases linearly from rest to 1.5× CO, but then stabilizes at 2× CO.

### 3.3 Comparison with FSI simulation

Flow waveforms acquired at the AAo from the HMCL experiments were used as the inlet boundary conditions for FSI simulations. The flow and pressure waveforms measured from the FSI simulations can be compared to those obtained in the HMCL. Figure 5 shows the waveforms at the AAo inlet in P-3 at 1.25× CO exercise. Although there is a generally good agreement between the waveforms measured using the two methods, a time delay is observed between the HMCL and FSI as we go downstream from the AAo. Additionally, while the flow splits to the branch vessels and the DAo in the FSI simulation are matched to the HMCL flow splits, there is a difference in the magnitude of the peak flows at these outlets.

**Fig. 5.**
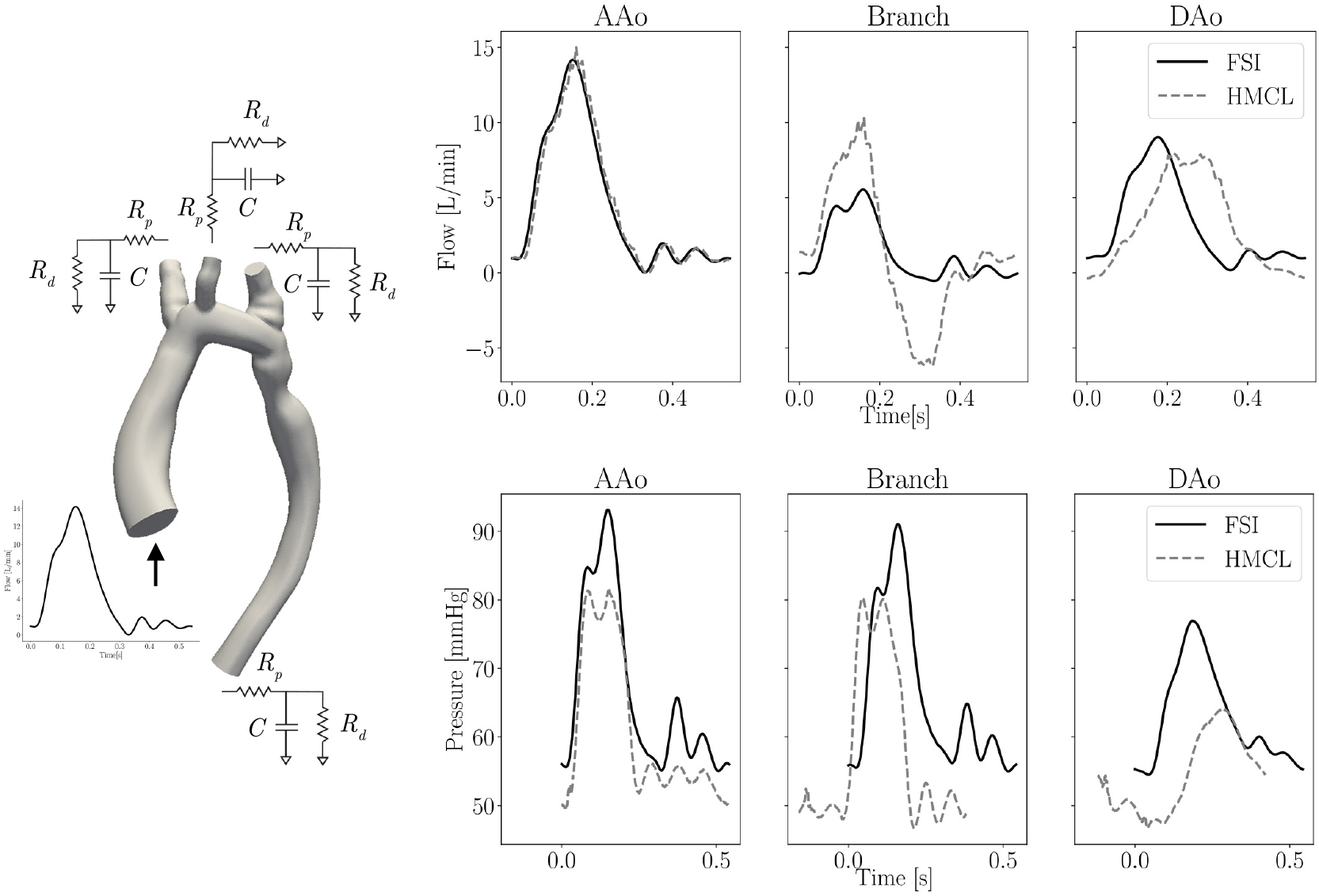
Schematic of boundary conditions used for FSI simulations (left). Comparison of flow (top) and pressure (bottom) waveforms acquired in the HMCL and via FSI simulations at three different locations - AAo inlet, head and neck vessels outlet (“Branch”), and DAo outlet. The waveforms from P-3 at 1.25× CO are depicted here as a representative example.

To compare the accuracy of FSI pressure measurements against those from the HMCL, the specific values of ΔP_FSI_ and ΔP_HMCL_ are provided in Table 3. A mean absolute error of 1.6 mmHg was observed between the two measurement methods with a standard deviation of 1.1 mmHg. The trend in mean ΔP with increasing CO for each patient using HMCL and FSI measurements can also be visualized in Figure 6A. There is good agreement in the trends observed in ΔP_HMCL_ and ΔP_FSI_. The discrepancy between ΔP_HMCL_ and ΔP_FSI_ in P-1 at 1.25× CO occurs from a physical limitation and is discussed later. The agreement between ΔP_HMCL_ and ΔP_FSI_ is further evaluated using Bland-Altman analysis, as shown in Figure 6B. ΔP_FSI_ always underestimated ΔP_HMCL_ but had a fairly tight confidence interval. The bias (mean of differences) was -1.6 mmHg, and the corresponding limits of agreement were [-0.5, -3.7] mmHg.

**Table 3.**
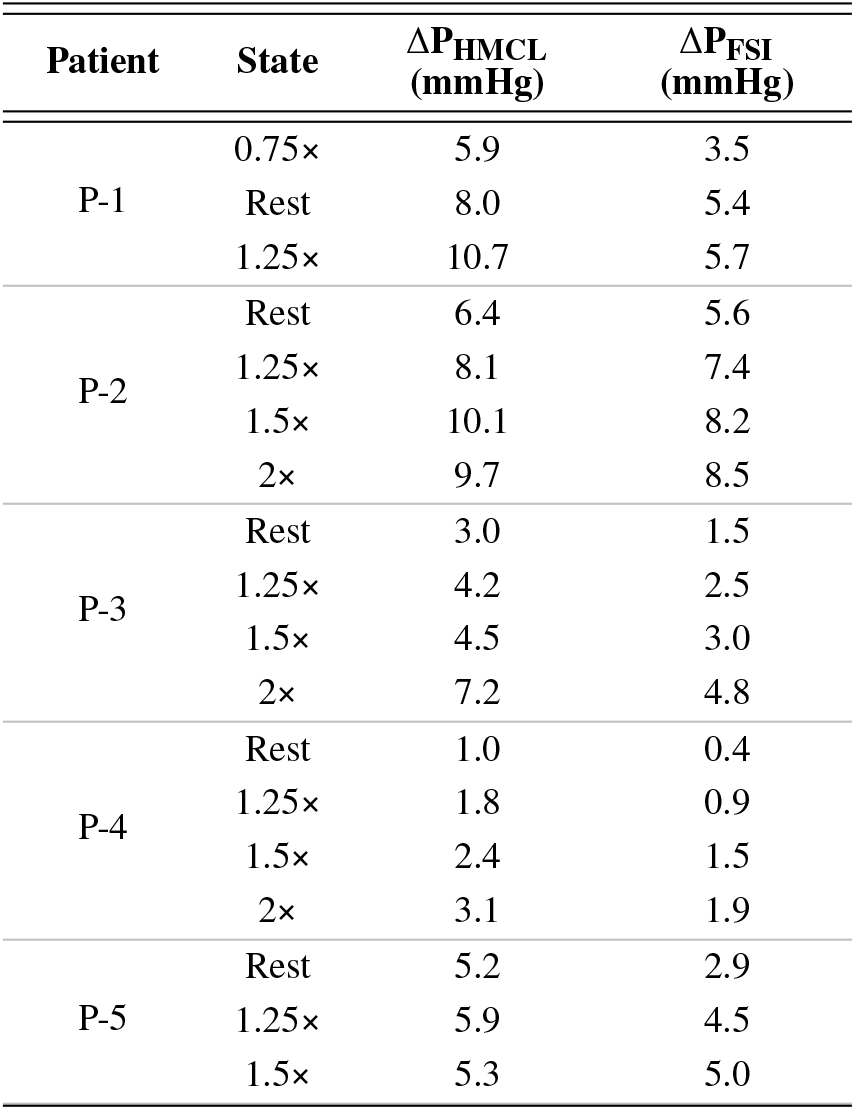
Mean ΔP between AAo and DAo in N=5 patients at rest and different states of exercise estimated from the HMCL (ΔP_HMCL_) and from FSI simulations (ΔP_FSI_).

**Fig. 6.**
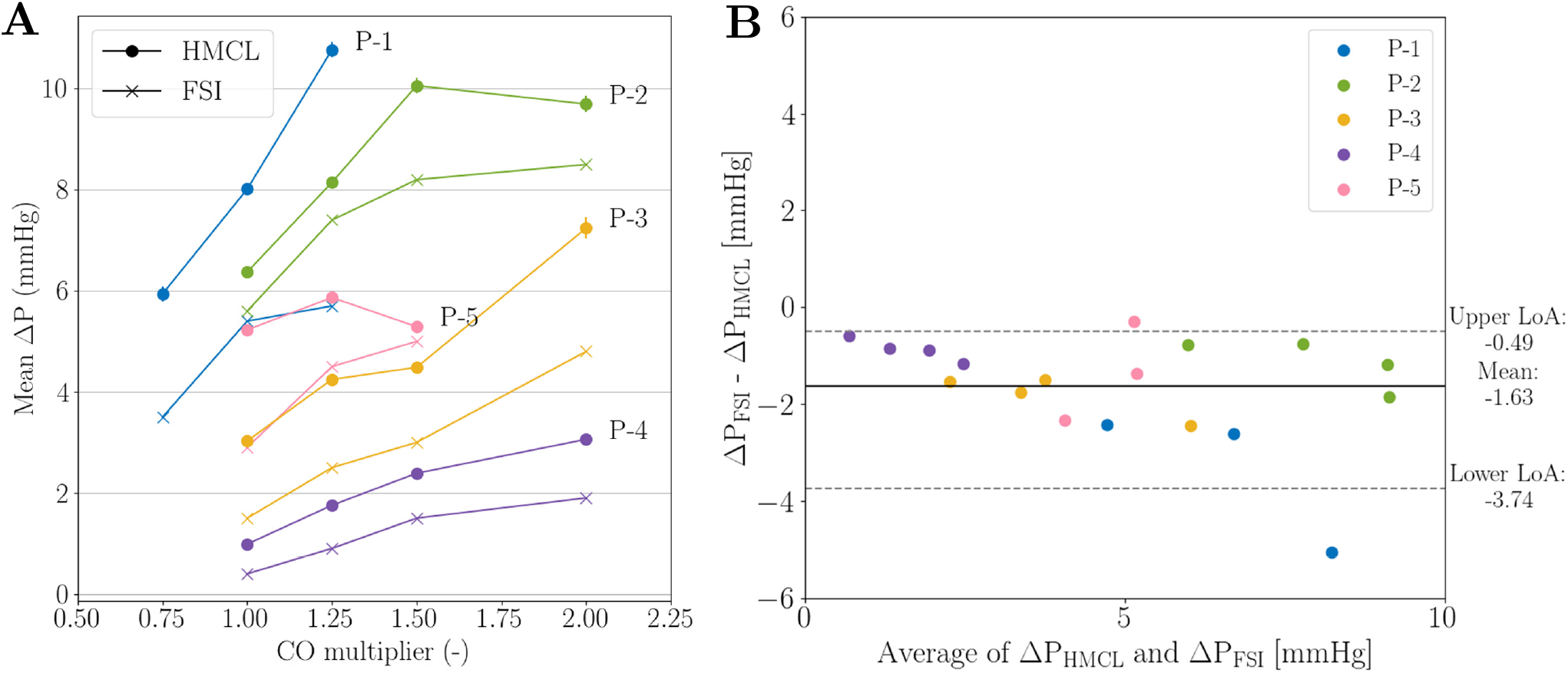
(A) Comparison of mean ΔP estimates from the HMCL and from FSI simulations at different hemodynamic states. (B) Bland-Altman plots of mean ΔP estimates from the HMCL and from FSI simulations. The solid line represents the mean difference between ΔP_HMCL_ and ΔP_FSI_, and the paired dotted lines correspond to the 95% limits of agreement (LoA).

## 4 Discussion

In current clinical practice, invasive cardiac catheterization is the gold-standard method used to assess the severity of CoA and make subsequent treatment decisions. However, the invasive nature of the procedure makes it expensive and can put the patient at increased risk for complications. We have previously demonstrated the ability to non-invasively estimate ΔP at rest using 0D and FSI simulations validated against invasive catheter measurements [22]. However, in the clinical setting, catheterization is only performed while patients are in an anesthetized state thereby likely underestimating the actual disease burden during day to day activities. This study addresses this gap and demonstrates through the use of an HMCL and FSI simulations that exercise-induced ΔP estimates cannot just be linearly extrapolated from ΔP measurements at rest in patients with CoA.

The HMCL provided a unique and versatile test-bed for assessing hemodynamics on the benchtop during exercise. It combined both *in vitro* and *in silico* components through incorporation of patient-specific compliant aortic phantoms and a closed-loop numerical representation of the cardiovascular system. The closedloop cardiovascular LPN allowed efficient tuning of the system to accurately recapitulate patient-specific flows and pressures on the benchtop at rest and exercise. The strength of this study was further enhanced through comparison of *in vitro* measurements with those from corresponding FSI simulations. A combined 0D-3D FSI simulations approach [22] was used to efficiently tune boundary conditions, initialize 3D simulations, and to compare to ΔP measurements from the HMCL. The excellent agreement in trends between the two methods validates our approach for studying exercise states that are difficult to obtain in current clinical practice. The patient-specific non-linear increase in the mean ΔP with increasing CO indicates the importance of predicting ΔP at exercise in patients with CoA. In a prior study combining MRI-ergometry with computational simulations, Schubert *et al*. demonstrated that CoA patients with normal ΔP at rest can have ΔP above the thresholds for intervention during exercise [4]. However, this was not true in all patients, and demonstrates the need for individualized assessment of the functional significance of CoA. These patients also exhibited an insignificant increase in stroke volume with exercise - the increase in cardiac index was mainly driven by an increase in HR, which is in line with our study. In a study performed by Mandell *et al*. in 3D-printed rigid phantoms of repaired CoA using 4D-Flow MRI, they demonstrated significant increases in ΔP, wall shear stress, vorticity and helicity with exercise. They also identified these secondary flow characteristics to be more strongly associated with exercise capacity than ΔP alone. LaDisa *et al*.’s *in silico* study demonstrated variations in exercise-induced ΔP increase between different CoA severity and different surgical intervention techniques [36]. They observed substantial increases in systolic BP and mean and peak BP gradients in patients with native CoA from rest to simulated moderate exercise. Peak BP gradients in treated CoA patients, however, were still higher than normal, indicating that simply restoring favorable anatomy through intervention may not always restore normal hemodynamics. All of the above studies support our finding about the patient-specific nature of the increase in ΔP with exercise. However, these studies focused on rest and only one state of exercise, making it impossible to assess the linearity of the increase. By assessing the hemodynamics in controlled conditions and at multiple intensities of exercise, we were able to establish the non-linearity in the trend of ΔP with increasing CO as well as observe variations in this trend between patients. This novel finding has important clinical implications.

Further, the pathophysiology of hypertension and exercise-induced hypertension in patients with CoA is complex and postulated to be due to a combination of endothelial dysfunction, abnormal arterial smooth muscle reactivity, changes in ventricular and vascular stiffness etc [37–42]. The results from our study therefore emphasize the need for further research into the factors that influence the non-linearity of ΔP and BP increase during exercise. Future studies may introduce inhomogeneous aortic wall stiffness or more physiologically driven tuning of the closed-loop cardiovascular LPN.

In addition to validating the non-linearity in ΔP increase, this study establishes FSI simulations as a useful tool to non-invasively and efficiently determine exercise hemodynamics in patients with CoA. Previous experimental studies, including our own, emphasize the difficulty of achieving exercise states with high flows, both *in vitro* and *in vivo*. These limitations can be easily overcome in computational simulations. Further, FSI simulations can be used to study more advanced flow indices such as wall shear stress and oscillatory shear index which can have significant clinical implications but cannot be measured clinically [4,36].

This study has several limitations. Only patients with CoA for whom 4D-Flow MRI as well as catheterization information was available were included in the study, thereby limiting the sample size. All the patient-specific aorta phantoms were 3D-printed with the same material and therefore have the same Young’s modulus, which may result in a mismatch with the patient’s native vessel wall properties. In addition, due to hardware limitations, we could not achieve instantaneous flow rates > 430 mL/s in the HMCL. As a result, the peak flows between the output from the closed-loop cardiovascular LPN connected to the HMCL and that measured in the HMCL are not well matched. Therefore, we report mean ΔP values instead of peak systolic ΔP. The phantom for P-5 burst beyond 1.5× CO, so no ΔP measurement at 2× CO is provided. Moreover, since P-1 had a high resting CO of ~8 L/min, we couldn’t achieve the 1.5× and 2× CO exercise states on the HMCL in this patient due to instabilities introduced in the cardiovascular LPN. Therefore, to evaluate the trend in mean ΔP in the patient, we acquired pressure measurements at 0.75× CO instead. Lastly, the blood volume calculated for the closed-loop cardiovascular LPN is estimated based on the patient’s weight [43]. Given that P-1 had a high resting CO, achieving even the 1.25× CO in P-1 on the HMCL was difficult and required manipulation of the parameters of the cardiovascular LPN likely beyond what is physiologically possible. This could explain the drop in mean BP going from rest to 1.25× CO, as well as the large discrepancy observed between ΔP_HMCL_ and ΔP_FSI_ for that data point.

The versatility of our approach is a notable advantage – HMCLs can be used to study hemodynamics for diagnosis and treatment in a variety of cardiovascular diseases as well as in different patient geometries and hemodynamic states. Future studies could expand the use of the HMCL to assess treatment outcomes in patients with CoA by assessing the impact of stent intervention or surgical corrections on aorta hemodynamics and better inform treatment decisions.

In conclusion, we have demonstrated the capability of using an HMCL compared against FSI simulations to comprehensively assess the severity of CoA over a range of cardiac outputs that are currently unattainable in the clinic and can guide patient-specific treatment planning.

## Data Availability

All simulation data from this work will be available on the Vascular Model Repository (vascularmodel.org)

## Acknowledgments

This work was funded by Digital Futures at KTH, VINNOVA (2022-00849), the Digitalisation platform at KTH, Stanford’s Maternal and Child Health Research Institute, and the American Scandinavian Foundation. Thank you to Dr. Marianne Schmid Daners at ETH Zurich for sharing her know-how in replicating the HMCL.

## Notes

### Competing Interest Statement

The authors have declared no competing interest.

### Author Declarations

IRB of Stanford University gave ethical approval for this work.

